# You Will Grasp Again: A Direct Spinal Cord/Computer Interface with the Spared Motor Neurons Restores the Dexterous Control of the Paralyzed Hand after Chronic Spinal Cord Injury

**DOI:** 10.1101/2022.09.09.22279611

**Authors:** Daniela Souza Oliveira, Matthias Ponfick, Dominik I. Braun, Marius Osswald, Marek Sierotowicz, Satyaki Chatterjee, Douglas Weber, Bjoern Eskofier, Claudio Castellini, Dario Farina, Thomas Mehari Kinfe, Alessandro Del Vecchio

## Abstract

The paralysis of the muscles controlling the hand dramatically limits the quality of life of individuals living with spinal cord injury (SCI). Here, we present a non-invasive neural interface technology that will change the lives of individuals living with cervical SCI (C4-C6). We demonstrate that eight motor- and sensory-complete SCI individuals (C5-C6, n = 7; C4, n = 1) are still able to task-modulate in real-time the activity of populations of spinal motor neurons with spared corticospinal pathways. In all tested patients, we identified groups of motor units under voluntary control that encoded a variety of hand movements. The motor unit discharges were mapped into more than 10 degrees of freedom, ranging from grasping to individual hand digit flexions and extensions. We then mapped the neural dynamics into a real-time controlled virtual hand. The patients were able to match the cue hand posture by proportionally controlling four degrees of freedom (opening and closing the hand and index flexion/extension). These results demonstrate that wearable muscle sensors provide access to voluntarily controlled neural activity in complete cervical SCI individuals.

---

Impaired hand function is arguably one of the most severe motor deficits in subjects with SCI, especially when bilateral^1^. There are currently no effective treatments for regaining control of the hand after muscle paralysis. Hand surgery is established, although not possible in every case, and with several limitations^2^. Restoration of hand function has so far been achieved by neural interfaces recording the activity of the motor cortex^3^, either through closed-loop electrical stimulation of the muscle^4^ or by controlling external devices^5^. However, besides the relatively poor control, invasive cortical implants are also an option limited to a small proportion of patients because of the surgical risks and long-term stability of the implant. Other neural interfaces involve the delivery of electrical stimulations in the spinal cord that indirectly targets the activity of the alpha motor neurons^6^.

The neural information most directly associated with behavior is the activity of spinal alpha motor neurons, which are the final common pathways of the neuromuscular system^7^. The activity of spinal motor neurons encodes movement through a simple linear transformation (the dynamics of the twitch forces of the muscle units) and therefore movement intent can be decoded directly. Almost all SCI are due to contusions of the spinal cord, which could leave some spared connections above and below the level of the injury^8^. While this spared neural activity is not sufficient to drive muscles for the generation of detectable forces, it can be used to infer motor intent and therefore to decode movements. Accordingly, we have recently reported in a single motor-complete SCI (C5-C6) individual, as a case study, the presence of a significant number of task-modulated motor units encoding the flexion and extension of individual fingers through a wearable, non-invasive neural interface^9^. That case study was a proof of concept in a single patient, and it was limited to offline analysis without any demonstration of patient-in-the-loop control. Here, we provide for the first-time evidence of voluntarily controlled spinal motor neurons in a relatively large group of SCI individuals (motor and sensory complete ranging from C4 to C6, Figure 1, Table 1, Video 1, 2). Through the decomposition of the high-density electromyogram (HDsEMG)^10–12^, we observed the presence of active motor neurons in all tested patients (Figure 1).

**Table 1.** Characteristics of research participants, including the average number of motor units (MUs) identified per task (mean ± SD) for each subject

| Subject | Age range (years) | Gender | Injury level | AIS | Wrist movement | Time since injury (years) | Number of MUs/task |
| --- | --- | --- | --- | --- | --- | --- | --- |
| S1 | 36-40 | M | C6 | B | yes | 18.8 | 14.5 ± 2.0 |
| S2 | 31-35 | M | C5 | B | yes | 9.1 | 8.1 ± 1.2 |
| S3 | 41-45 | F | C6 | B | yes | 24.2 | 3.5 ± 1.4 |
| S4 | 36-40 | F | C5 | A | yes | 24.2 | 7.3 ± 2.3 |
| S5 | 31-35 | M | C4 | A | no | 22.2 | 8.4 ± 0.7 |
| S6 | 56-60 | M | C5 | A | no | 6.9 | 22.8 ± 4.2 |
| S7 | 41-45 | M | C6 | C | no | 18.2 | 7.4 ± 2.0 |
| S8 | 36-40 | F | C5 | B | yes | 5.0 | 5.9 ± 1.2 |

**Figure 1.**
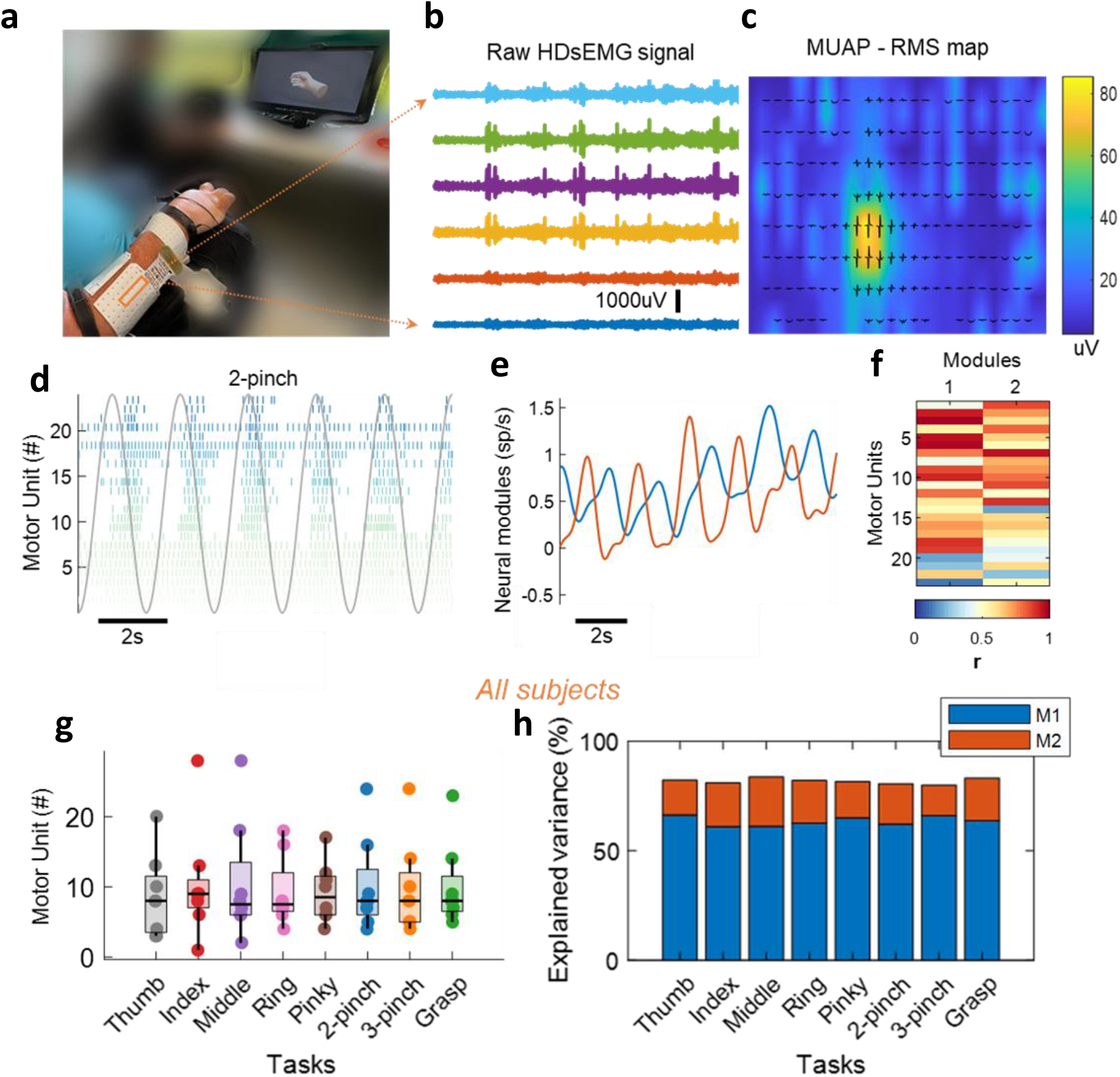
**a**. Experimental setup consisting of 320 surface EMG electrodes placed in the forearm muscles. The movement instructions were guided by a virtual hand video displayed on a monitor in front of the subject. **b**. A few example electrodes show raw HDsEMG signals while the subject attempts a grasp task (flexion and extension of the fingers, 0.5Hz). **c**. Example of spatial mapping based on the root mean square values of the motor unit action potential. **d**. Raster plot of motor unit firings (color-coded) identified during 10s of the two-finger pinch task. **e**. Neural modules extracted for the same task, using factorization analysis. **f**. Pearson correlation values (r) of the individual motor units with the two neural modules. **g**. Number of identified motor units (MUs) across all tasks and subjects (each dot represents one subject). **h**. Percentage of explained variance by the two neural modules (M1 – blue and M2 – red) averaged across all subjects for each task.

Figure 1 shows an overview of the offline experiments where subjects were asked to match the visual cue displayed through a virtual hand. The virtual hand displayed hand opening and closing, two and three-finger pinch, and individual digits movement (flexion and extension) at 0.5Hz movement velocity. Figure 1a shows the experimental setup, with 320 electrodes placed on the proximal and distal forearm muscles and tendons. Figure 1b shows six EMG channels and a motor unit waveform superimposed on a heatmap based on the root mean square activity (Figure 1c). In all tested patients, we observed clear motor unit action potentials with high signal-to-noise ratios (>30dB^13^). We then looked at how these motor units were controlled by studying the association between motor unit activation times (Figure 1d) and the movement trajectories of the digit tip of the virtual hand (grey lines in 1d). The raster plot in Figure 1d shows a clear grouping of motor units encoding flexion and extension movements during the two-finger pinch task. As in our previous experiment^9^, we used a factorization method to retrieve the motor dimension (flexion and extension of the motor units, 1e-f).

For all tested individuals, we were able to consistently identify some motor neurons that were controlling the flexion and extension movements (Supplementary Figs.). Figure 1g-h shows a summary of all subjects and tasks. For all the tasks (Fig. 1g-h), we identified a specific subpopulation of motor units that encoded that specific movement, with an average of 9.8 ± 0.7 motor units per task. Because of the large number of units, we were able to identify unique units virtually in all recorded tasks, which gives a perfect classification accuracy for all these motor dimensions. Therefore, after years of cervical spinal cord injuries leading to motor and sensory complete paralysis (ranging from 5.0 to 24.2 years, Table 1), these subjects still had spared connections from motor cortex impinging the activity of spinal motor neurons. This is evidenced by the fact that these motor units showed high voluntary modulation that matched with high degrees of accuracy the virtual hand movements (Fig. 2a). Figure 2a shows all the identified motor units for two individuals and all tasks. These previous results are based on the prediction accuracies and number of motor dimensions from the offline decomposition of the HDsEMG.

**Figure 2.**
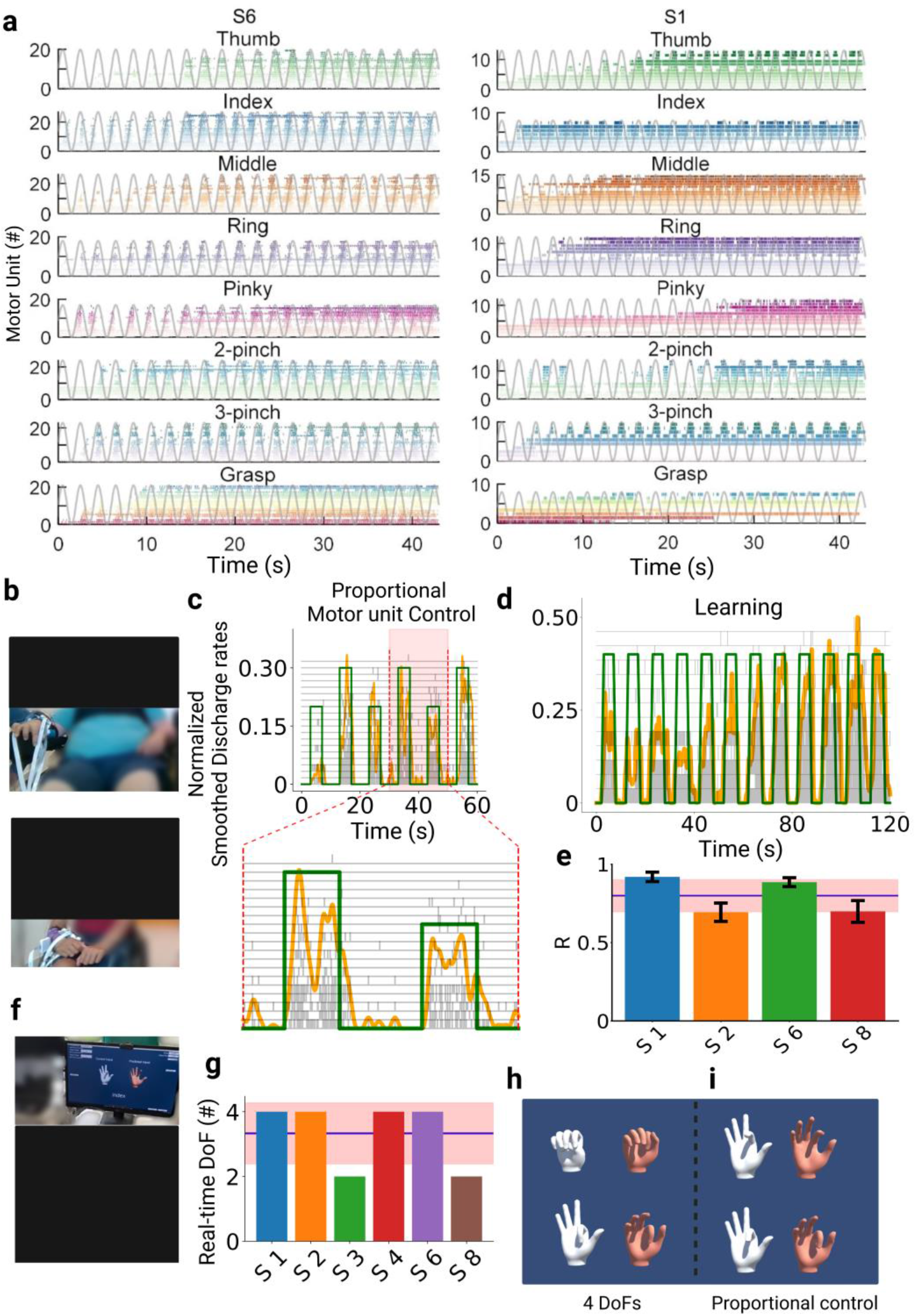
**a**. Raster plot for all motor units identified during the respective task (color-coded) and the virtual hand movement trajectories (grey line). Note the task-modulated activity of the motor unit firing patterns, that encoded flexion and extension movements. **b**. Real-time tasks for two participants (S1 and S6). **c**. The participants were asked to follow a trajectory on a screen (green line) by imagining a grasp movement. The motor unit were decomposed online, and the cumulative smoothed discharge rate (yellow line) was used as biofeedback. After few seconds of training (**d**), the subjects could track the trajectories with very high accuracies and at different target levels (**c**). **e**. Cross-correlation coefficient (R) between the smoothed discharge rate and the requested tasks for 4 subjects. **f**. After the online motor unit decomposition, we used a supervised machine learning method to proportionally control the movement of a virtual hand. Four out of six subjects were able to proportionally open and close the hand (**g-i**), and proportionally control in both movement directions (flexion and extension) the index finger (**h-i**). These subjects were able to control four degrees of freedom (DoFs) that corresponded to hand opening, closing, index flexion and extension.

In a second experiment, which was collected on average 3-5 months after the first session, six subjects were tested again with a similar experimental procedure but tuned for real-time control. The subjects were asked to proportionally control a moving cursor on a screen based on the real-time decoding of the discharge timings of motor neurons (Figure 2c-d). Moreover, the tested individuals also controlled a virtual hand (Figure 2f-i, Video 2), demonstrating full voluntary control of the decoded neural activity.

We developed a real-time mapping of the discharge timings of motor neurons so that the patients could control a virtual hand and a cursor on the screen with the motor unit discharge activity and the HDsEMG signal (Figure 2, Video 1). After a few seconds of training (see Fig. 2d), the subjects were able to control the motor unit firing patterns and progressive recruitment of motor units at different target forces and with high accuracies (Fig. 2b-d). In this experiment, we also used a supervised machine learning algorithm to control a virtual hand (Fig. 2f-i, Video 2).

Video 1 shows a subject that controls the activity of groups of motor units in real-time, modulating the recruitment and discharge rate to proportionally hit two different target levels of activation. The motor neuron discharge times were summed and normalized in real time to the number of active neurons so that the patients could modulate a moving object (yellow cursor, Fig. 2c-d) by increasing/decreasing the discharge rates. Figure 2c shows the proportional control of two target levels that was mediated by both the concurrent recruitment of additional units (grey raster plot) and higher discharge rates. Figure 2d shows a complete recording set that lasted 120 seconds. Note that just after 50 seconds of training the subject was able to move the cursor on relatively high levels of normalized motor unit activity. The scaling of the motor unit activity is based on a simple equation that considers the maximal motor unit discharge activity and the highest number of motor units that were identified during an off-line calibration trial, that lasted 10 seconds for each trained task.

We then trained the subjects to move a virtual hand that was displayed on a monitor and to match the movement of a control hand (Fig. 2f-i, Video 2). After this training, the subjects were able to proportionally open and close the hand with high levels of accuracy, when compared to the control hand instructions (Fig. 2i, Video 2). Most of the subjects were able to proportionally flex and extend the index finger (two degrees of freedom) and open and close the hand (two degrees of freedom). Figure 2f shows the subject’s view: the monitor displayed two hands, a control hand (white color), and a second hand that was controlled by a regression-based machine learning algorithm. Four out of 6 subjects (Fig. 2g) were able to control four degrees of freedom consisting of proportional control of index flexion and extension and hand opening and closing (Fig. 2h-i, Video 2). It is important to note that each experiment across all patients did not last more than 3 hours, with most of the time used for placing the electrodes and explaining the tasks. Although we did not measure the time it took for the subject to control the virtual hand and 2D cursor control, it surely did not take more than 30 minutes accounting even for the subjects with the highest level of wrist and hand paralysis. This can be further improved once the subjects are trained with the task. In sum, the presented technology has a direct clinical translation for both home and hospital use for restoring and monitoring the spared corticospinal connections after traumatic SCI.

The results presented above provide for the first-time evidence of voluntarily controlled spinal motor neurons in a relatively large group of SCI subjects (motor and sensory complete ranging from C4 to C6) that have been paralyzed for decades. We observed the presence of active modulation of motor neuron activity in all tested patients. We then developed a real-time mapping of the discharge timings of motoneurons so that the patients could control a virtual hand. The tested patients performed virtual hand tasks accurately and proportionally, demonstrating full voluntary control of the decoded neural activity. The results indicate that motor- and sensory-complete SCI patients maintain relevant neural activity as output of the spinal cord circuits below the lesion and that they can accurately control this activity to regain hand function. Wearable muscle sensors are therefore a technology that may compete in terms of clinical viability and efficacy with invasive brain or spine implants for restoring hand function in complete SCI patients. We contend that the proposed non-invasive approach is a clinically superior solution to hand function restoration in SCI than the current invasive brain and spinal neural interfaces.

## Supporting information

Video 1 - Online Decomposition - Subject 6

Video 2 - Virtual Hand Control - Subject 6

## Data Availability

All data produced in the present study are available upon reasonable request to the authors.

## Methods

Eight participants with spinal cord injury (SCI) were recruited for this study (Seven individuals with chronic motor complete SCI and one with motor incomplete SCI). The inclusion criteria were: (1) injury level C4-C6 and (2) age between 18 and 60 years old (3) absence of voluntary movement of one hand or both hands. Participants S6 and S7 presented movement of the left hand. All participants gave their written informed consent to take part in the study. The study was conducted in agreement with the Declaration of Helsinki, and it was approved by the Friedrich-Alexander-Universität Ethics Committee (application 22-138-Bm).

### Study overview/Experimental protocol

This study was conducted in two sessions. In the first session, subjects were instructed to attempt movements shown by videos of a virtual hand while high-density electromyographic (HDsEMG) signals from their forearm were recorded. For the second session, 6 subjects returned after 3-5 months of the first session, in which a regression model (based on global EMG) and/or an online decomposition method was used to decode movement intention, according to their HDsEMG signals.

In the first session, 320 HDsEMG electrodes were placed in the forearm of the participants’ dominant hand (subject 7 was paralyzed only on the non-dominant hand). The subjects were then asked to stay in a comfortable position with their arm (Fig. 1a and 2b,f). With a computer monitor in front of them, videos of a virtual hand performing different tasks were displayed and the participants were instructed to attempt the movements accordingly. The tasks included movement of the individual digits, grasp, two-finger pinch, and three-finger pinch at two different speeds (0.5Hz and 1.5Hz), and lasted 42s each. Two trials were performed for each movement. Only data from 0.5Hz movements were analyzed due to the difficulty of the subjects in performing fast movements.

In the second session, using the same electrode configuration as the first visit, first, an EMG-to-activation regression model was built: subjects were asked to attempt a full flexion of their fingers according to the tasks that were easier for them to perform, and their EMG signals were acquired and associated to synthetic ground truth representing maximal activation for the relevant degrees of freedom. After that, the participants attempted the flexion/extension of the digits, and the predicted activation was shown to them in real time through a virtual hand interface (‘predicted hand’). A virtual hand showing a predefined movement (referred here as ‘control hand’, Fig. 2f,h-i) was used to help the subjects to perform the movements and for further analysis.

Also in this session, we tested a real-time EMG decomposition approach (offline decomposition followed by online decomposition). We used 128 HDsEMG electrodes to assess if the subjects would be able to follow a digital trajectory with their motor units smoothed cumulative discharge rate. First, during the offline decomposition, HDsEMG data were recorded while the participants were asked to attempt a maximum flexion of the digits (10s per task). The recorded data is decomposed as described in the ‘Online decomposition’ section and the decomposition results are stored for the online task.

Subsequently, in the online decomposition step, the subjects were instructed to follow a periodic rectangular waveform trajectory shown on a monitor, with 10s period (5s of rest in between), for 60-120s. For two consecutive periods, the subjects were asked to attempt flexion and extension of the same digits as performed in the offline decomposition. The motor unit firings detected with this method (smoothed motor unit firings) were also shown as feedback to the subjects.

Lastly, we also tested if the subjects would be able to increase their discharge rate and progressive recruitment of motor units by increasing the height of the ramp. The rectangular trajectories have two different activation levels, 20% and 30% of maximum neural activation. The discharge rate was normalized using the maximum discharge rate obtained during the brief offline decomposition step.

### Virtual hand videos and interface

The 3D model of the hand used to instruct the subjects during the recordings was developed in Blender (Blender 3.0, Blender Foundation). The appearance of the virtual hand was modified to resemble a real human hand, a texture was applied to provide a better experience to the participants. Thirteen videos of the hand model were generated in Blender, each of them corresponding to one task/hand movement (individual digits flexion/extension at 0.5 Hz and 1.5Hz, grasp, two-finger pinch, and three-finger pinch). Figure 1a shows an example of the virtual hand that was displayed to the subjects.

The virtual hand interface was created to communicate with the regression model. This interface was generated using the software Unity (Unity Software Inc). An improved virtual hand model (mesh object from VIVE Wave SDK - https://hub.vive.com/storage/docs/en-us/index.html) was used, which allows more degrees of freedom for the right hand (with 17 bones). The interface shows two virtual hands: the former shows the movements to be performed (control hand) and the latter (predicted hand) is connected to the regression model and shows the output obtained directly from the HDsEMG signals. Therefore, by comparing the control and the predicted hand, we can understand the amount of flexibility of control that is spared after the injury.

### HDsEMG recordings

For electrode placement, the forearm skin was shaved and cleansed with 70% ethyl alcohol. The ulna bone was marked with a skin marker. Five grids of 64 surface EMG electrodes were placed over the forearm muscles (3 squared grids of 8 rows x 8 columns configuration, with interelectrode distance (IED) of 10mm; 2 rectangular grids - 13×5, IED = 8mm; OT Bioelettronica, Turin, Italy). The squared grids were placed aligned to the ulna bone, while the rectangular ones were placed posterior and anterior in the forearm, above the wrist joint. The grids were attached to the skin using bi-adhesive foam, that was placed aligned to the electrodes and filled with conductive paste (SpesMedica, Battipaglia, Italy). The grids were then secured with tape. The reference for the electrode grids was placed on the elbow joint, and the main ground electrode was placed on the styloid process of the ulna. The HDsEMG signals were recorded using a multichannel amplifier 16-bit A/D (Quattrocento, OT Bioelettronica). The signals were recorded in monopolar mode using the software OT BioLab+, with sampling frequency of 2048Hz and a bandpass filter 10-500Hz. The recordings were synchronized with the start of the virtual hand videos. For the second part of the experiments, HDsEMG signals were streamed in real-time using a Transmission Control Protocol/Internet Protocol (TCP/IP) for communication. For this, the software OT BioLab Light was used, with the following settings: 2048Hz sampling frequency, bandpass filter 10-500Hz, and 8Hz refresh rate (for online decomposition) or 16Hz refresh (for the regression model).

### Online decomposition

To decompose the EMG signal into individual motor units, we used a combination of fast independent component analysis (fastICA) and convolutive kernel compensation (CKC). This method involves a convolutive sphering (extension and whitening) of the measurement matrix, followed by an iterative optimization of separation vectors that maximizes the non-gaussianity. This approach allows an automatic decomposition of the sources. Video 1 shows the real-time decomposition and the modulated motor unit activity for a subject with SCI and Figure 2c-e the performance of the subjects to track a line on a screen by modulating the discharge rates of the motor units.

A script in Python was developed to show the motor unit firings detected with this method (smoothed motor unit firings) as feedback to the subjects, while they were requested to follow a rectangular trajectory of 10s period (Video1 and Figure 2c-e). A cross-correlation was applied between the target levels and the normalized cumulative discharge rates (Figure 2c). The average correlation coefficient of every 20s trials for each subject was used as a metric.

The online decomposition is divided into two parts. The first part is the offline decomposition, in which the EMG data is recorded for each task the subject was asked to perform in real-time. The length of the recorded signal is 10s, and the subjects were asked to perform a full flexion of one or more digits during the recording (in separate trials). Then, the recorded data is decomposed using fastICA, which is based on the convolutive blind source separation method (described below in detail). The results of fastICA are the individual extracted sources and their respective separation vector. The separation matrix and the individual action potentials of the motor units (MUAPs) are stored for the online task. The individual MUAPs are computed by the spike-triggered average (STA) using the extracted sources. Before the real-time decomposition begins, the individual separation matrices are merged into a single matrix, and duplicates are flagged by calculating the Pearson correlation coefficient between the MUAPs.

During real-time decomposition, a periodic rectangular waveform trajectory (10s period, 5s of rest in between) is displayed continuously to the subject for one to two minutes. The waveforms have two different required activation levels with 20% and 30% of maximum neural activation. For each period, the subject was asked to perform one of the tasks from the offline decomposition. To circumvent the computational complexity obstacle of whitening in real-time, the observations from the same controlled task are expanded and then directly multiplied by the stored separation matrix from the offline part. The results of this multiplication are considered as the extracted sources and then correlated with the MUAP templates (template matching). To distinguish between noise and neural activity, the maximum noise power is calculated in the first data frame where the subject is assumed not to move (no ramp). If the calculated correlation coefficient is above a predefined threshold and the signal has an SNR higher than a scalar multiple of the maximum noise level (see below), it is classified as a spike.

To decompose the HDsEMG signal into the individual motor units in real-time, the general approach of offline decomposition followed by online decomposition described by Barsakcioglu and Farina (2018) was adopted^14^. The offline decomposition method is based upon the iterative extraction of sources from the convolutive sphering (extension and whitening) of observations described in Negro et al. (2016)^15^ and Holobar and Zazula^16^. We have tailored the current general structure of the algorithm for the current data from SCI individuals, as described below.

We can model the HDsEMG generation process as a convolutive mixture of the pulse trains and the action potentials of motor units, in matrix form:

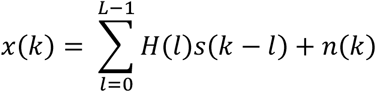

where *x*(*k*) = [*x*_1_(*k*), *x*_2_(*k*), *x*_3_(*k*), …, *x*_*m*_(*k*)]^*T*^ are the HDsEMG signals recorded from *m* EMG channels (number of observations) with *x*_*i*_(*k*) being the signal recorded from the *i*^*th*^ channel, *s*(*k*) = [*s*_1_(*k*), *s*_2_(*k*), *s*_3_(*k*), …, *s*_*n*_(*k*)]^*T*^ are the firing patterns of *n* motor units (number of sources). *H* is the mixing matrix of dimension *m × n* which carries the information of motor unit action potentials (MUAP), with *L* being the during of the action potentials and *l* each sample; *n(k)* is the unidentified noise term for each channel. With the motivation of increasing the ratio between the number of observations and the number of sources, the observations are extended using R-lagged samples, where R=1000/m^15^. From the extended observations, a whitening matrix is derived by performing eigenvalue decomposition of the covariance matrix of the extended observations. Then a fixed-point iterative algorithm with Gram-Schmidt Orthogonalization is used to maximize the number of uniquely identified sources. Next, a silhouette score-based K-means driven approach is adopted to detect the spikes from the decomposed source that involves the second iteration of the CKC approach to remove the unreliable sources, as described in^15^. This offline decomposition prior to online decomposition can be conceptualized as a training phase from which the separation matrix is extracted which contains the separation vectors. Also, the MUAP templates from the extracted sources are generated using spike-triggered-averaging (STA) from the offline decomposition and stored in a matrix.

In the real-time (online decomposition phase), the observations obtained from the same controlled task are extended and then directly multiplied with the separation matrix to avoid the impediment of computational complexity due to whitening. The outcomes of this multiplication are considered to be extracted sources and then subjected to a further 2D cross-correlation template matching technique where the MUAPs extracted for each of the sources were used as the template. Since the MUAPs are the results of the convolution of the action potential and Dirac delta pulses, we can obtain MUAP shapes from their discharge times as having a repetition of the action potential at the time instances of the Dirac pulses. Therefore, we used the shapes of the action potentials to perform a cross-correlation between each of the samples of the extracted single MUAP and the MUAP of the corresponding channels in real-time, for a further validation step. The Pearson’s normalized cross-correlation coefficient is defined as,

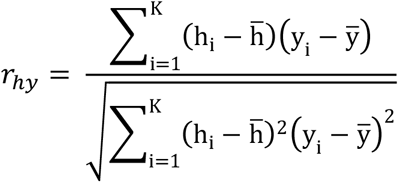

where, *h* is the template, i.e. the MUAP shape, and *y* is the windowed segment of size *K* of the single MUAP which has to be correlated to the MUAP shape to detect the spikes. This window is rolled over the entire signal to find the correlation coefficient concerning the MUAP. From the outcome of the first frame of the real-time sEMG decomposition, the maximum noise power was also calculated because the first frame in our experimental setup contained no physical activity. The samples in the extracted sources of the real-time decomposition which showed a cross-correlation coefficient more than a predefined threshold and exhibit SNR higher than a scalar multiple times the maximum noise level was sorted as a spike. This approach allows an automatic decomposition of the sources.

The spike sorting in the offline decomposition task was performed using the Silhouette threshold of 0.9. In order to set up the real-time decomposition, a range of cross-correlation coefficients between 0.50-0.70 was chosen to perform the template matching-based spike sorting. To perform the template matching, MUAPs of the length of 30 samples, i.e. approximately 15ms long MUAPs were extracted through spike-triggered averaging using the spike time instances and the decomposed SMUAPs. The performance of the real-time EMG decomposition was evaluated both qualitatively, i.e. through visualization of the overall shape of the envelope of the firings, and quantitatively by computing the rate of agreement (ROA). The ROA is defined as:

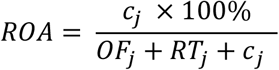

Where *c*_*j*_ is the total number of firings of the *j*^*th*^ motor unit identified by both offline and online decomposition, *RT*_*j*_ is equal to the total number of firings of the *j*^*th*^ motor unit identified in online decomposition only, *OF*_*j*_ is the total number of firings of the *j*^*th*^ motor unit identified by offline decomposition only. The choice of the hyperparameters *r*_*hy*_ and SNR threshold required to perform spike-sorting was made from an isometric ramp task experiment with 64 channel HD-sEMG signal collected from the tibialis anterior muscle of a healthy individual by picking up the combination yielding the best ROA obtained by performing online decomposition. The correlation coefficient of 0.60 and SNR threshold of 10 was selected to set up the real-time decomposition framework as they indicated high accuracy between offline and online decomposition^14^.

### Regression-based simultaneous and proportional control

A regression-based machine-learning method was used to simultaneously and proportionally estimate the amount of required activation of each degree of freedom of the virtual hand. Namely, we used Incremental Ridge Regression ^17^. Ground truth was provided by artificially associating minimal and maximal activation values to EMG signals gathered from the participants while they were being asked to minimally and maximally perform specific actions (i.e., flexion and extension of the digits)^18^. The bandpass filtered EMG signals from the Quattrocento were processed as follows: first, a non-overlapping moving window FIR low pass filter was applied (cut-off frequency 200Hz). In order to improve visualization, the signal was multiplied by a factor of 35. Then, the absolute value of the signal was extracted. Then, the signal was low-pass filtered again through a first-order IIR filter with a response of the form *y*(*k*) = *αx*(*k*) + (1 − *α*)*y*(*k* − 1) (cutoff frequency 1Hz). After this, an adaptive filter was employed. This is a first-order IIR filter, the cutoff frequency *F*_*c*_ of which is determined by the instantaneous value of the signal *s*(*t*) itself and of its first-order time derivative according to *F*_*c*_= exp(8|*IIR*_*α*=0.6_(*x*(*k*) − *x*(*k*−1))|−7 | *x*(*k*)|), where 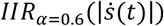 represents the first-order time derivative of the signal low pass filtered through an IIR filter with *α* = 0.6. The signal was then subsampled at 25Hz. An adaptive IIR filter like the one described above was also applied to the prediction from the ridge regression model. The values for all the coefficients used for signal filtering were set empirically following a series of pre-tests.

Despite its simplicity, Ridge Regression has recently been proven highly effective when applied to the problem of multi-DoF simultaneous and proportional prosthetic myocontrol using high-density *surface* electromyography^19–21^. The adaptive filter is used here to exploit the heteroscedasticity of the EMG signal, which tends to be noisier if the mean value is higher.

### Offline HDsEMG decomposition

The monopolar HDsEMG signals were first band-pass filtered (20-500Hz) with a Butterworth second-order filter. The data from all 5 electrode grids were concatenated into one matrix. To identify the individual motor units, these HDsEMG signals were decomposed through a blind source separation method (convolutive kernel compensation algorithm). The software DEMUSE (v. 4.5; The University of Maribor, Slovenia) was used for that^16^, which automatically detects the motor unit discharge times. After that, a visual inspection of the identified motor unit spike trains was performed, to account for false positive/false negative detected spikes, as described previously^22^. Only the motor units with a pulse-to-noise ratio > 26dB before manual inspection were kept.

### Factor analysis

A dimension-reduction technique, factor analysis, was applied to the smoothed discharge rate of all motor units identified for a task. The discharge rate was first low-pass filtered with a Hann window of 1s (1Hz). Afterward, the function *factoran* from Matlab was applied to the matrix with the smoothed discharge rates without any orthogonality constraint. This function uses the maximum likelihood method to estimate the common factors associated to the observed data (motor units discharge times). The Pearson correlation between the motor units’ discharge rate and the fitted factors was then calculated. The observed data are assumed to be based on a linear combination of the common factors that explain most of the data variance. The percentage of the total variance explained by these factors was also calculated for each task from each subject and later averaged across all subjects for each one of the tasks.

