## Supplementary Information for "You Will Grasp Again: A Direct Spinal Cord/Computer Interface with the Spared Motor Neurons Restores the Dexterous Control of the Paralyzed Hand after Chronic Spinal Cord Injury"

- Video 1: https://www.youtube.com/watch?v=__17_VkVhLU

**Video 1** | Electromyography (EMG) online decomposition. The video shows a participant with spinal cord injury (Subject 6) following a trajectory on a screen (blue line on the top of the screen) using the cumulative discharge rate (red line on top of the screen), shown as biofeedback. The target level of the trajectory is set at 20% and 30% of the maximal estimated neural drive, which is estimated during the offline decomposition step considering the maximum discharge rate.
The lower part of the screen shows in real-time the discharge rate of the motor units that were identified during the offline decomposition step.

- Video 2: https://www.youtube.com/watch?v=LwYQjNXv63o

**Video 2** | Virtual hand control in real-time. The video shows the virtual hand interface integrated with a regression-based machine learning method that allows the proportional control of the movement of a virtual hand using electromyography (EMG) activity. The control hand is the one instructing the movement to be performed. The predicted hand is connected to the regression model and shows the output obtained from the EMG signals. Note that the subject with spinal cord injury (S6) is able to proportionally control the movement of the index finger (flexion and extension) and to open and close the hand. In total, the subject was able to control four degrees of freedom.

- One figure for each of the patients, showing all motor units by task, and MRI spinal cord (8 Figures)

# S1

M, C6, AIS B

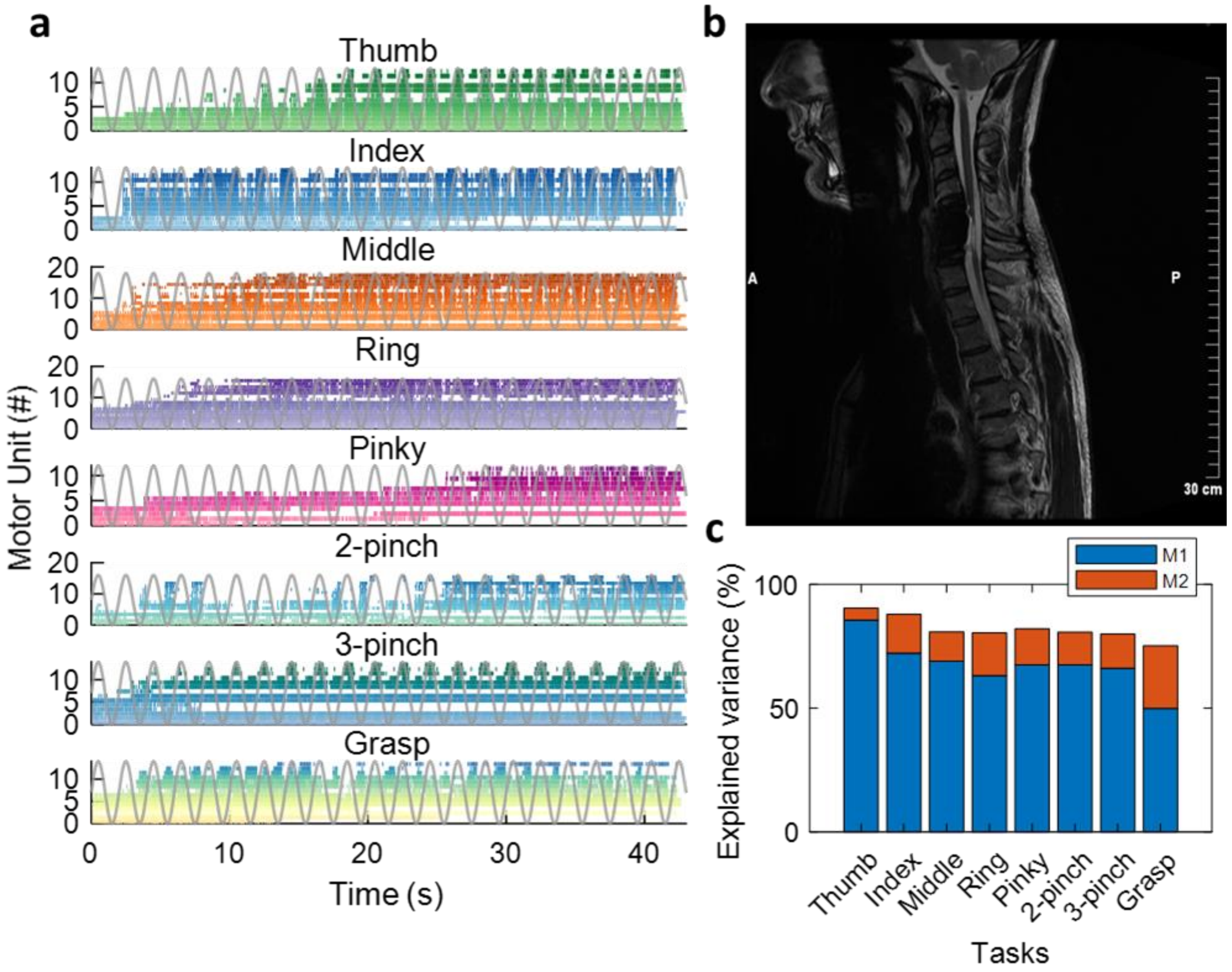


**Supplementary Figure 1** | Data from Subject 1 (S1): male with C6 injury and AIS B (ASIA impairment scale) **a.** Raster plot for all motor units identified during the respective task (color-coded) and the virtual hand movement trajectories, 0.5 Hz frequency (grey line). Note the task-modulated activity of the motor unit firing patterns, that encoded flexion and extension movements. **b.** Sagittal T2-weighted magnetic resonance image (MRI) of cervical and thoracic spine, image from patient's medical history. **c.** Percentage of explained variance by the two neural modules (M1 - blue and M2 - red) for each task.

# S2

M, C5, AIS B

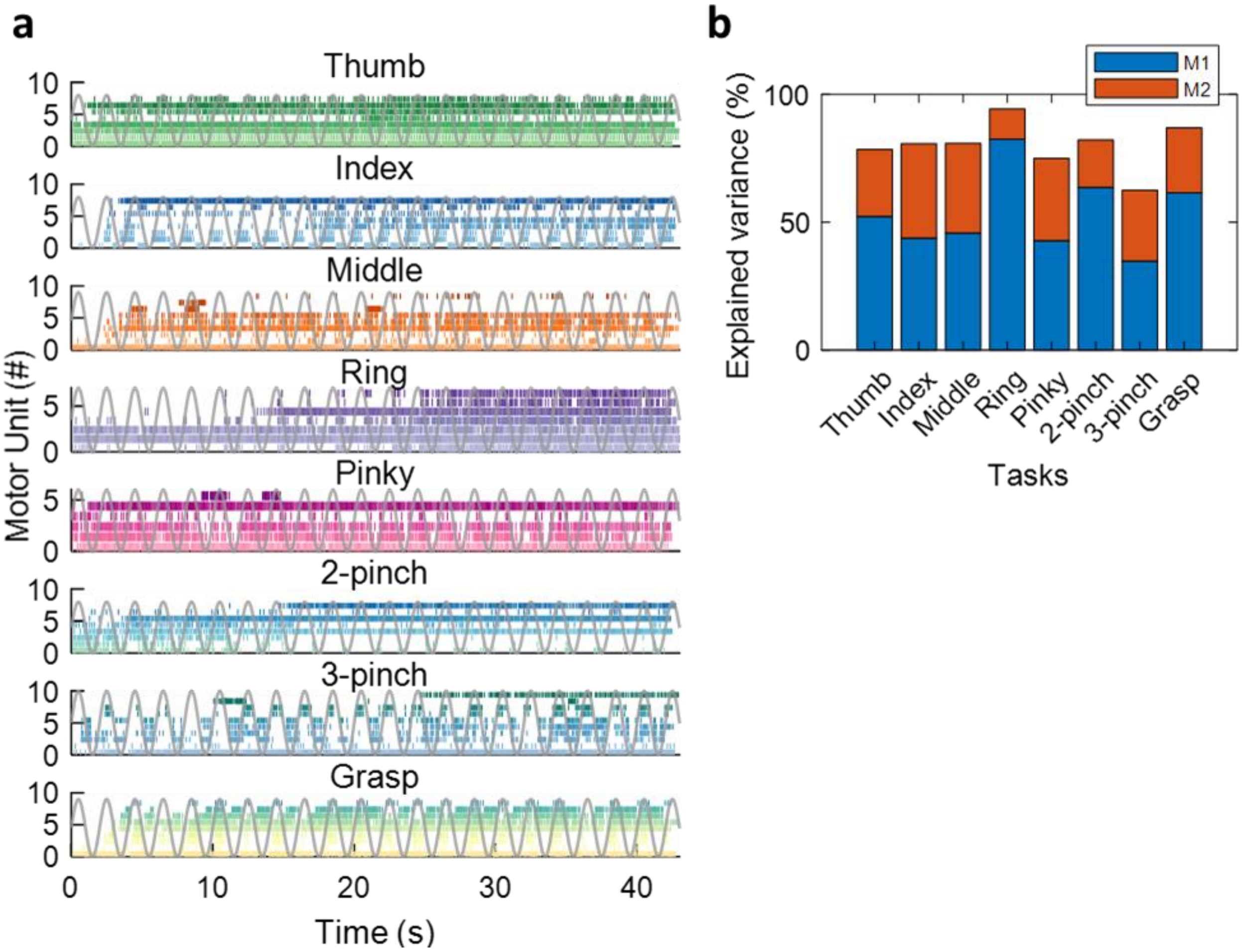


**Supplementary Figure 2** | Data from Subject 2 (S2): male with C5 injury and AIS B (ASIA impairment scale) **a.** Raster plot for all motor units identified during the respective task (color-coded) and the virtual hand movement trajectories, 0.5 Hz frequency (grey line). Note the task-modulated activity of the motor unit firing patterns, that encoded flexion and extension movements. **b.** Percentage of explained variance by the two neural modules (M1 - blue and M2 - red) for each task.

# S3

F, C6, AIS B

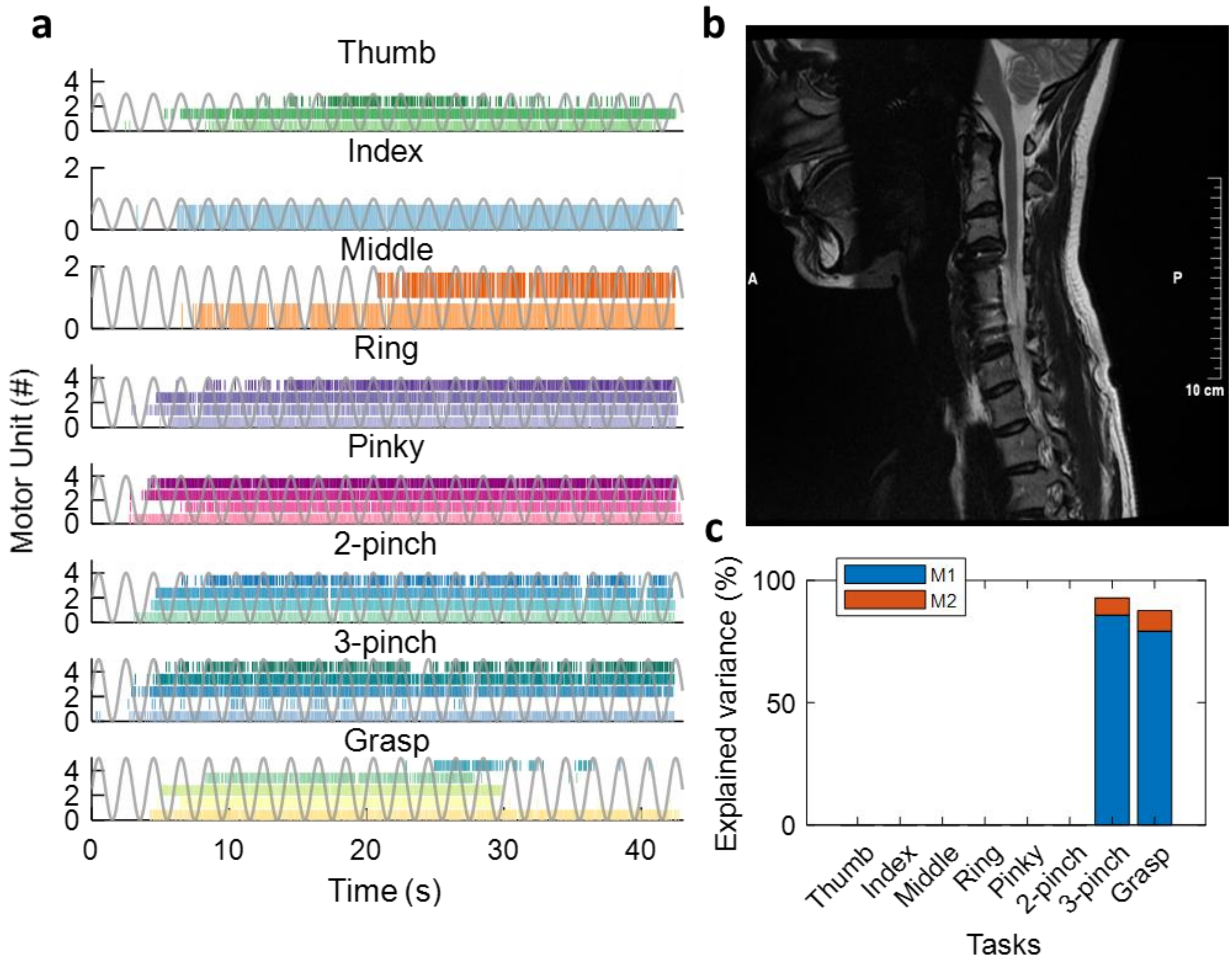


**Supplementary Figure 3** | Data from Subject 3 (S3): female with C6 injury and AIS B (ASIA impairment scale) **a.** Raster plot for all motor units identified during the respective task (color-coded) and the virtual hand movement trajectories, 0.5 Hz frequency (grey line). Note the task-modulated activity of the motor unit firing patterns, that encoded flexion and extension movements. **b.** Sagittal T2-weighted magnetic resonance image (MRI) of cervical spine, image from patient's medical history. **c.** Percentage of explained variance by the two neural modules (M1 - blue and M2 - red) for each task. Only three-finger pinch and grasp tasks presented enough motor units to extract the neural modules.

# S4

F, C5, AIS A

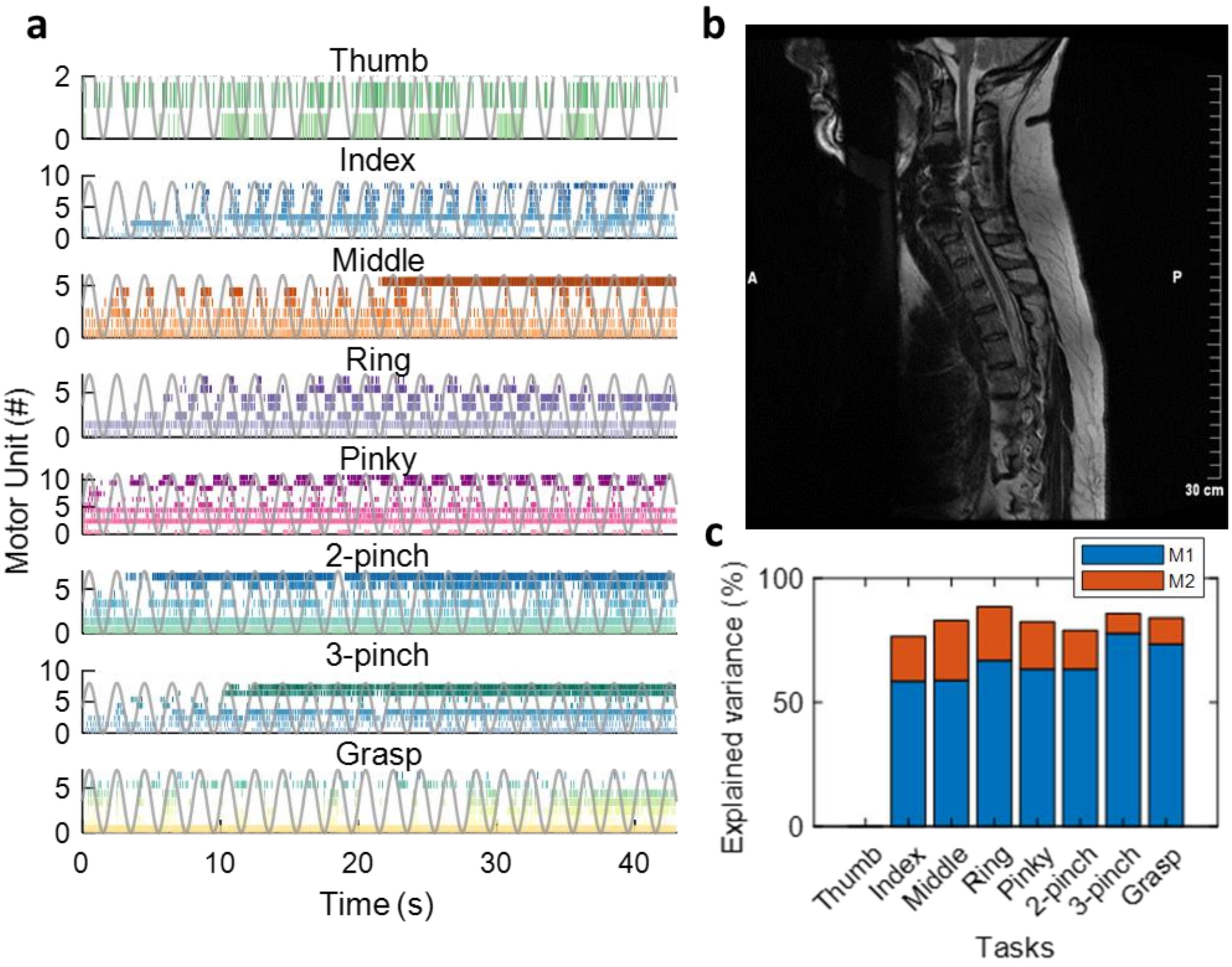


**Supplementary Figure 4** | Data from Subject 4 (S4): female with C5 injury and AIS A (ASIA impairment scale) **a.** Raster plot for all motor units identified during the respective task (color-coded) and the virtual hand movement trajectories, 0.5 Hz frequency (grey line). Note the task-modulated activity of the motor unit firing patterns, that encoded flexion and extension movements. **b.** Sagittal T2-weighted magnetic resonance image (MRI) of cervical and thoracic spine, image from patient's medical history. **c.** Percentage of explained variance by the two neural modules (M1 - blue and M2 - red) for each task. Thumb task did not present enough motor units to extract the neural modules.

# S5

M, C4, AIS A

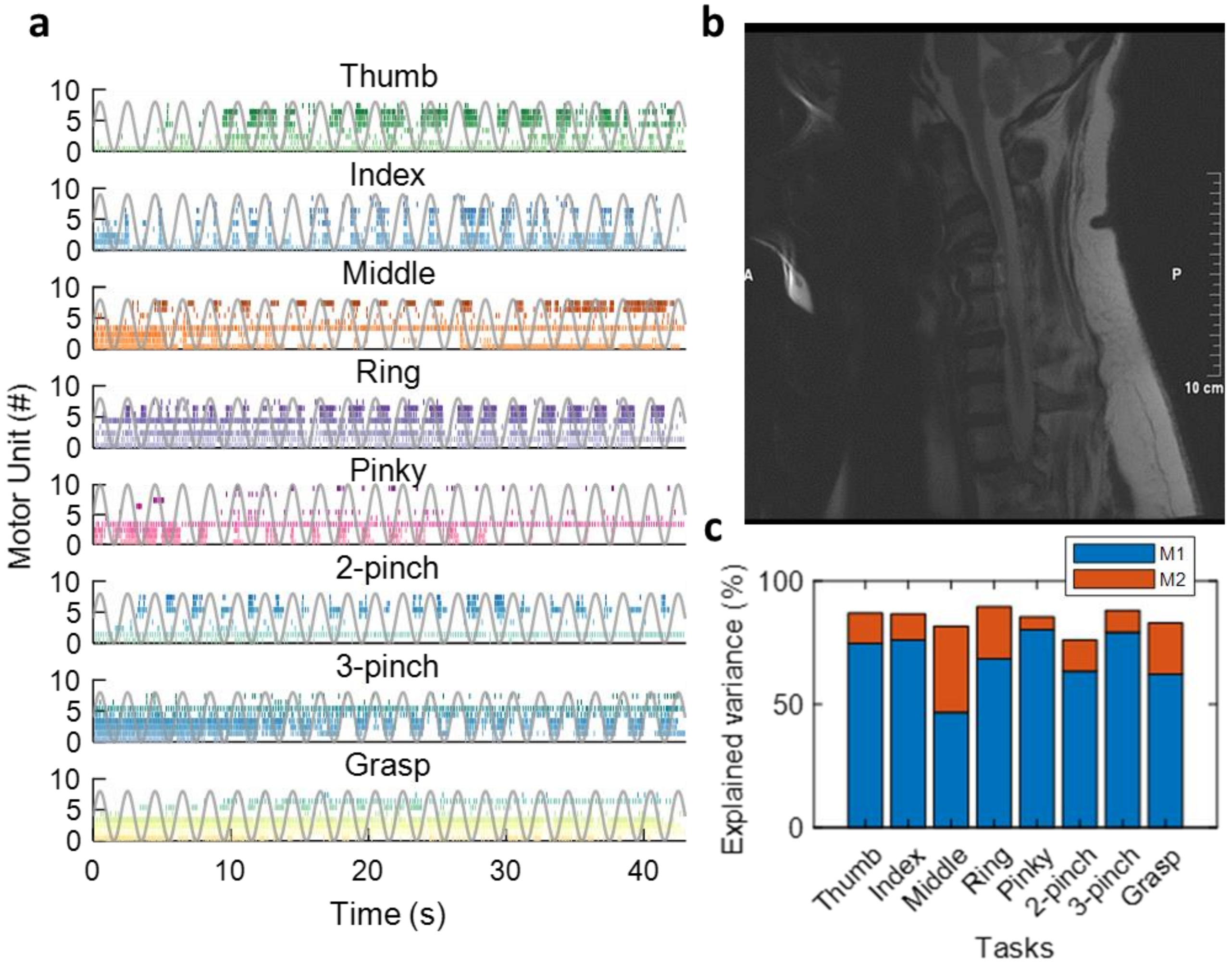


**Supplementary Figure 5** | Data from Subject 5 (S5): male with C4 injury and AIS A (ASIA impairment scale) **a.** Raster plot for all motor units identified during the respective task (color-coded) and the virtual hand movement trajectories, 0.5 Hz frequency (grey line). Note the task-modulated activity of the motor unit firing patterns, that encoded flexion and extension movements. **b.** Sagittal T2-weighted magnetic resonance image (MRI) of cervical spine, image from patient's medical history. **c.** Percentage of explained variance by the two neural modules (M1 - blue and M2 - red) for each task.

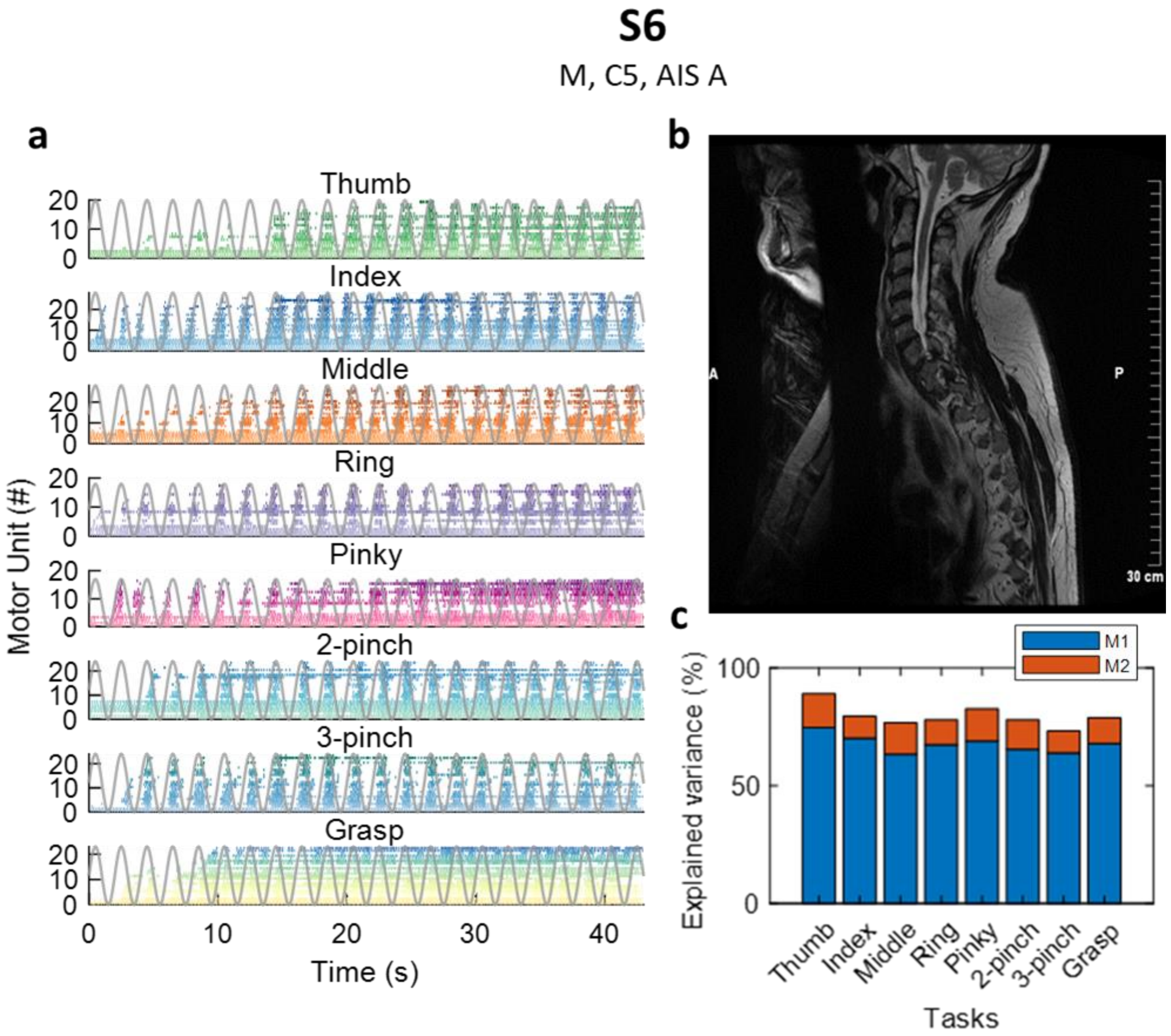


**Supplementary Figure 6** | Data from Subject 6 (S6): male with C5 injury and AIS A (ASIA impairment scale) **a.** Raster plot for all motor units identified during the respective task (color-coded) and the virtual hand movement trajectories, 0.5 Hz frequency (grey line). Note the task-modulated activity of the motor unit firing patterns, that encoded flexion and extension movements. **b.** Sagittal T2-weighted magnetic resonance image (MRI) of cervical and thoracic spine, image from patient's medical history. **c.** Percentage of explained variance by the two neural modules (M1 - blue and M2 - red) for each task.

**S7**

M, C6, AIS C

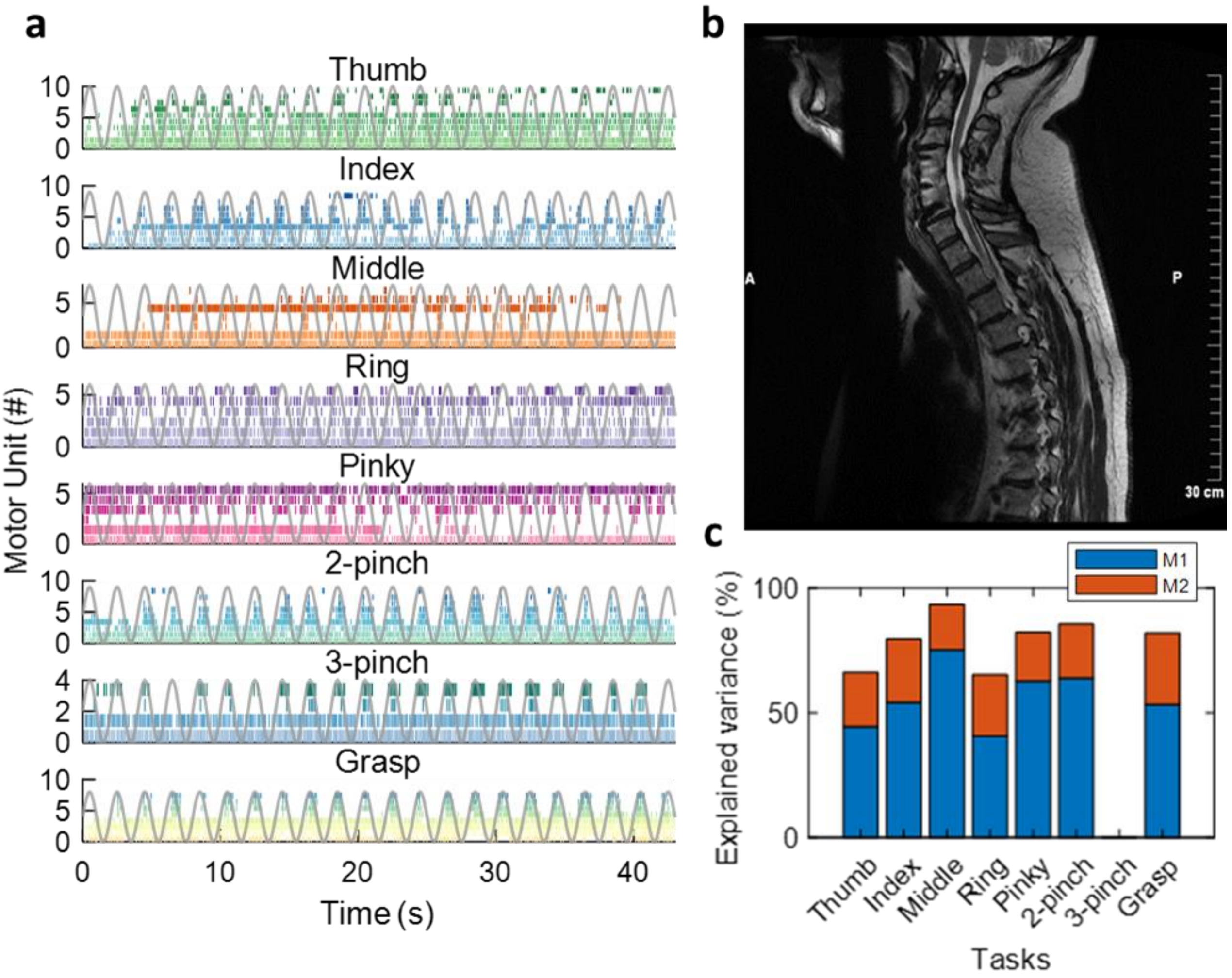


**Supplementary Figure 7** | Data from Subject 7 (S7): male with C6 injury and AIS C (ASIA impairment scale) **a.** Raster plot for all motor units identified during the respective task (color-coded) and the virtual hand movement trajectories, 0.5 Hz frequency (grey line). Note the task-modulated activity of the motor unit firing patterns, that encoded flexion and extension movements. **b.** Sagittal T2-weighted magnetic resonance image (MRI) of cervical and thoracic spine, image from patient's medical history. **c.** Percentage of explained variance by the two neural modules (M1 - blue and M2 - red) for each task. Three-finger pinch task did not present enough motor units to extract the neural modules.

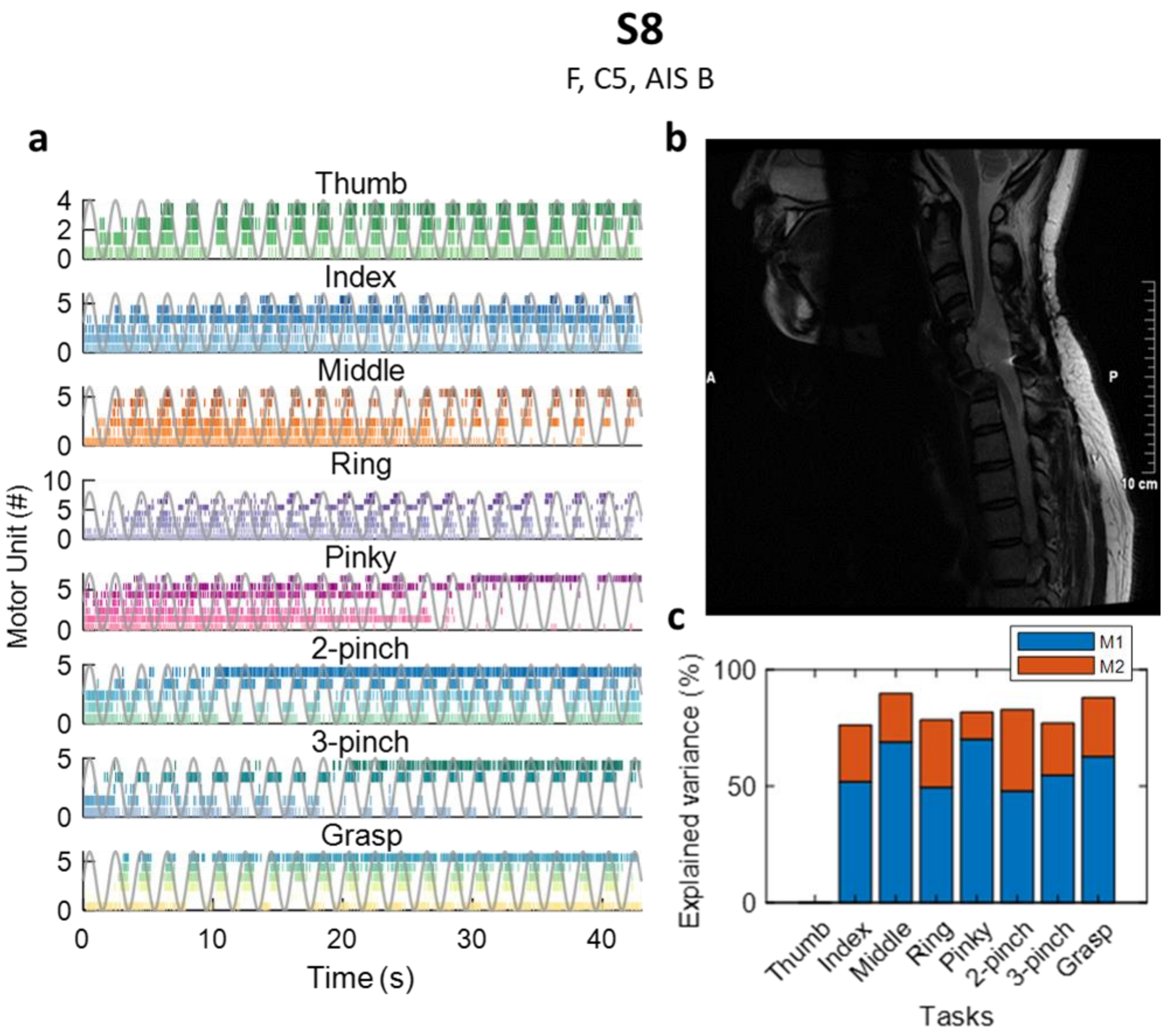


**Supplementary Figure 8** | Data from Subject 8 (S8): female with C5 injury and AIS B (ASIA impairment scale) **a.** Raster plot for all motor units identified during the respective task (color-coded) and the virtual hand movement trajectories, 0.5 Hz frequency (grey line). Note the task-modulated activity of the motor unit firing patterns, that encoded flexion and extension movements. **b.** Sagittal T2-weighted magnetic resonance image (MRI) of cervical spine, image from patient's medical history. **c.** Percentage of explained variance by the two neural modules (M1 - blue and M2 - red) for each task. Thumb task did not present enough motor units to extract the neural modules.
